# Data-Driven Early Prediction of Cerebral Palsy Using AutoML and interpretable kinematic features

**DOI:** 10.1101/2025.02.10.25322007

**Authors:** Melanie Segado, Laura Prosser, Andrea F. Duncan, Michelle J. Johnson, Konrad P. Kording

## Abstract

Early identification of cerebral palsy (CP) remains a major challenge due to the reliance on expert assessments that are time-intensive and not scalable. Consequently, a range of studies have aimed at using machine learning to predict CP scores based on motion tracking,e.g. from video data. These studies generally predict clinical scores which are a proxy for CP risk. However, clinicians do not REALLY want to estimate scores, they want to estimate the patients’ risk of developing clinical symptoms. Here we present a data-driven machine-learning (ML) pipeline that extracts movement features from infant video based motion tracking and estimates CP risk using AutoML. Using AutoSklearn, our framework minimizes risk of overfitting by abstracting away researcher-driver hyperparameter optimization. Trained on movement data from 3- to 4-month-old infants, our classifier predicts a highly indicative clinical score (General Movements Assessment [GMA]) with an ROC-AUC of 0.78 on a held-out test set, indicating that kinematic movement features capture clinically relevant variability. Without retraining, the same model predicts the risk of cerebral palsy outcomes at later clinical follow-ups with an ROC-AUC of 0.74, demonstrating that early motor representations generalize to long-term neurodevelopmental risk. We employ pre-registered lock-box validation to ensure rigorous performance evaluation. This study highlights the potential of AutoML-powered movement analytics for neurodevelopmental screening, demonstrating that data-driven feature extraction from movement trajectories can provide an interpretable and scalable approach to early risk assessment. By integrating pre-trained vision transformers, AutoML-driven model selection, and rigorous validation protocols, this work advances the use of video-derived movement features for scalable, data-driven clinical assessment, demonstrating how computational methods based on readily available data like infant videos can enhance early risk detection in neurodevelopmental disorders.

**CCS Concepts:**

- **Computing methodologies** → **Machine learning approaches**;
- **Applied computing** → *Health informatics*.

**ACM Reference Format:** Melanie Segado, Laura Prosser, Andrea F. Duncan, Michelle J. Johnson, and Konrad P. Kording.. Data-Driven Early Prediction of Cerebral Palsy Using AutoML and interpretable kinematic features. In. ACM, New York, NY, USA, 8 pages.

## 1 Introduction

Cerebral Palsy (CP) is a neuromotor disorder that results in significant physical disability, affecting approximately 1 in 300 children worldwide [30]. CP is often associated with perinatal complications, particularly hypoxia during birth, which can lead to early brain injury and subsequent motor impairments [32]. However, clinical evaluations such as the General Movement Assessment (GMA) that predict risk are typically only conducted for infants with known perinatal complications [13]. This means that many children without obvious early risk factors may go undiagnosed until significant motor delays emerge [32]. Moreover, the traditional diagnostic pathways rely on expert-driven assessments that are labor-intensive, non-scalable, and prone to variability. There is therefore a clear need for automated, data-driven approaches that can efficiently analyze movement patterns and reliably predict which children are at risk of developing CP.

Machine learning models have shown promise in predicting early indicators of CP, often through classification of clinical scores such as GMA [1, 18, 19, 22, 26, 31, 46]. There is increasing interest in camera-based approaches due to their low cost and wide-spread availability [10, 29, 34, 38, 45, 46]. However, these models typically rely on hand-engineered features and dataset-specific tuning, limiting their ability to generalize across populations despite showing promising results. Furthermore, predicting GMA score specifically rather than direct outcomes introduces an intermediate layer of uncertainty, reducing the model’s real-world applicability [48]. While our previous work demonstrated the feasibility of using AutoML to predict GMA scores [43], it did not address they key question of whether the trained model generalized to predicting CP diagnoses. To address this, we extend our prior methodology by directly evaluating whether early movement features retain predictive power for CP diagnoses at later clinical follow-ups.

**Figure 1:**
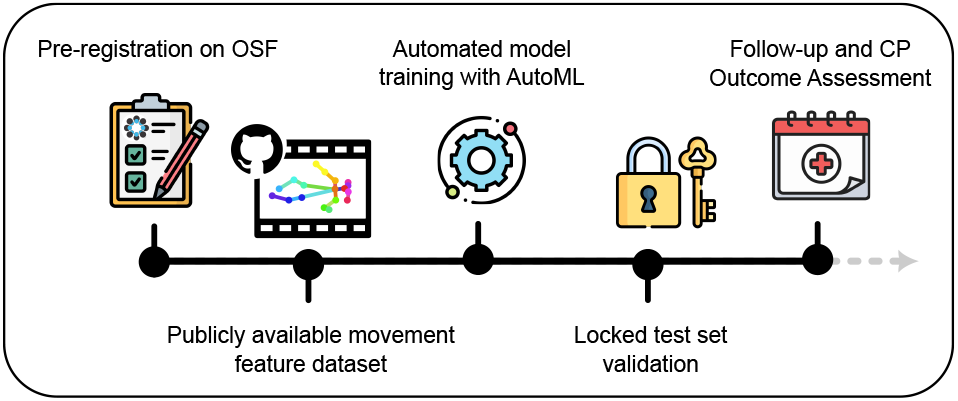
Pre-Registered Machine Learning Pipeline for CP Outcome Prediction. The study follows a structured, preregistered workflow to ensure transparency and reproducibility. First, all model parameters, features, and evaluation criteria were pre-registered on OSF. Movement data was collected and fully de-identified movement features were made publicly available on OSF, enabling independent validation. Automated machine learning (AutoML) was then used to extract features and train models for GMA score prediction, and code was made available on GitHub. A locked test set was used for unbiased validation, ensuring that no hyperparameter tuning influenced final performance. Finally, longitudinal follow-up at 2+ years allowed for CP outcome assessment, demonstrating that the model trained on early movement features generalizes to predicting CP diagnoses without retraining. This pipeline highlights the potential for scalable, data-driven clinical decision support while adhering to rigorous open science principles.

Our data-driven approach integrates video-based movement analysis with automated machine learning (AutoML) to determine whether CP can be predicted from movement features using rigorous methods that minimize the risk of overfitting 1. Specifically, we use Auto-Sklearn to automate feature selection, hyperparameter optimization, and model selection, to minimize the risk reduce the risks of overfitting [16, 17]. Unlike traditional methods that require dataset-specific model tuning, the Auto-Sklearn framework abstracts away manual optimization, reducing the risk of overfitting and increasing scalability [21]. We used a pre-trained vision transformer [52] and open source software packages for pose estimation [8] to obtain precise skeletal tracking from monocular handheld video, from which a kinematic feature set capturing kinematic complexity, symmetry, and temporal dynamics can be computed. By systematically integrating auto-sklearn with video-derived movement analytics enabled by open source tools, this work establishes a scalable, reproducible framework for neurodevelopmental screening. Our findings suggest that computational methods perform similarly to hand-tuned models in predicting long-term clinical outcomes, paving the way for automated, interpretable risk assessment in early childhood development. This study highlights the potential of AutoML-powered movement analytics to transform neurodevelopmental diagnostics by reducing reliance on expert assessments and improving early detection of CP in diverse, real-world settings. To evaluate whether a model trained to predict GMA scores can generalize to directly predict cerebral palsy outcomes, we conducted a longitudinal follow-up study through the neonatal follow-up program at CHOP. Among the 1,053 infants assessed at 3-4 months using the GMA, approximately 143 were later diagnosed with CP. We found that, without any retraining, the model initially optimized for GMA score prediction maintained good performance when applied to CP outcome prediction. This result underscores the effectiveness of methods optimized for generalizability, demonstrating that early movement features capture clinically meaningful neurodevelopmental risk signals. All analyses were conducted using the pre-registered lock-box validation set, ensuring rigorous performance evaluation.

## 2 Methods

Ethical approval for this study was provided by the University of Pennsylvania (Penn) Institutional Review Board (IRB Protocol Number: 833180), acting as the single IRB or record and a subsequent reliance agreement between Penn and the Children’s Hospital of Philadelphia (CHOP) Institutional Review Board (IRB Protocol Number: 19-016641). An IRB waiver was granted to assess the clinical outcomes of the Early Detection Trial (EDT) GMA cohort (IRB protocol number: 24-022210)

### 2.1 Video-Based Motion Tracking and Feature Computation

Video recordings of infant movement were collected using iPad devices and securely uploaded to a Redcap database, ensuring compliance with ethics guidelines and stored and analyzed within the CHOP hospital system. Access to this dataset was restricted to hospital staff and the Cerebral Palsy Foundation. Each recording captured a 1-2 minute video of the infant positioned supine in their crib, minimizing external distractions. To standardize assessments, infants were observed wearing minimal attire, with no pacifiers, toys, or external engagement, allowing for unobstructed tracking of spontaneous movements. See [43] for detailed dataset description. For pose estimation we use ViTPose-H, a computer vision foundation model fine-tuned for pose tracking on adults [11, 52], which we found generalized to infant pose tracking without the need for additional fine tuning. We normalized the resulting 2D skeletal time series with a mean and median filter to reduce noise (See: [6, 7, 42] for full methods description and code). From these, we computer 38 clinician-selected kinematic movement features (pre-registered in 2017) using publicly available Python code [41]. These features capture posture, velocity, acceleration, left-right symmetry, and complexity of movement and were averaged over 2-second sliding windows (1-second overlap) to preserve temporal information. We also computed A grand average across all frames. A full implementation of the pose estimation pipeline and feature extraction code is available on Zenodo [41], and the extracted movement feature dataset is publicly accessible on the OSF pre-registration site [40]. The movement complexity feature was computed as the Shannon entropy on the velocity time series of each joint, given by

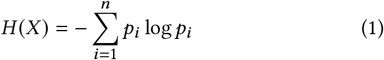

where *p*_*i*_ represents the probability distribution of discretized velocity magnitudes across time [44, 49]. Higher entropy values indicate greater variability in movement, while lower values suggest stereotyped or constrained motor patterns.

For consistency with previous machine learning models predicting GMA scores, only the global feature averages were used for model training and evaluation. Although prior analyses indicated that some features were more predictive of GMA scores than others, no additional feature selection was applied to avoid overfitting. The identical 38-feature pre-registered movement vector was used for both GMA prediction and CP outcome prediction, ensuring methodological consistency and enabling direct assessment of model generalization.

### 2.2 Model selection and hyperparameter optimization using Auto-sklearn

We trained an auto-sklearn model to predict GMA scores using “vanilla” settings specifically designed to minimize the risk of over-fitting [17]. These included limiting the model to an ensemble size of 1, avoiding the use of metalearning for initialization of paramters, and not allowing string features. It is generally not advisable to try to understand which models are selected by autoML algorithms; while predictions can be very good, the difference between distinct modeling choices often, maybe typically, would not be statistically significant. However, our test-readers all asked us what modeling choices are made by auto-sklearn, so here goes. The best model selected using these training constraints was a Stochastic Gradient Descent (SGD) classifier [51] with a modified Huber loss function, optimized using an inverse scaling learning rate schedule and L2 regularization (λ = 4.629 × 10^−5^) [35]. The Modified Huber Loss selected by auto-sklearn optimized is a robust loss function that blends properties of hinge loss and squared loss to improve classification performance in the presence of noisy labels. Unlike standard hinge loss used in support vector machines, Modified Huber Loss softens the margin constraint, making it more resilient to label noise while still promoting confident classification. This function provides quadratic penalization for small errors, encouraging smooth optimization, while linear penalization for large errors reduces sensitivity to outliers.The learning rate was initialized at η0 = 0.0169, and the epsilon parameter for the Huber loss was set at 7.437 × 10^−5^. Training proceeded for 64 iterations, with the weight update rule given by *w*_*t*+1_ = *w*_*t*_ −*η* (*∇L*(*w*_*t*_) +λ*w*_*t*_), where the learning rate follows an inverse scaling schedule defined as *η* = *η*_0_ /(1+t)^0.5^. To improve convergence, the model applied standardization to all numerical features using (*x* ^′^= (*x* − *μ*)/ *σ*). Missing values were imputed using mean substitution, and AutoML selected instance weighting to address class imbalance, adjusting class probabilities by (*w*_*c*_ = *N*/(|*C*| × *n*_*c*_)), where ( *N*) is the total sample size, (|*C*|) is the number of classes, and *n*_*c*_) is the number of samples in class (*c*). The final model was chosen based on validation loss minimization. The model achieved a cost function score of 0.319, reinforcing the effectiveness of automated hyperparameter selection in movement-based neurodevelopmental risk prediction. The procedure balances sensitivity to small classification errors with robustness against extreme misclassifications. CP diagnoses involve diagnostic uncertainty and inter-rater variability, and the selected loss function effectively handles label noise while ensuring model stability. This makes it particularly well-suited for movement-based neurodevelopmental risk prediction, where misclassified cases can arise due to subtle variations in early motor features. Key though is that all these choices are taken automatically by autoML without risk of overfitting or leakage.

### 2.3 Reproducibility and Model Validation

To ensure the reproducibility of GMA score prediction and account for updates to key Python libraries [20, 33, 50], we retrained the auto-sklearn classifier using the features released on the OSF pre-registration site. The training set was selected based on pre-registered IDs, and the model was trained using the same initial parameters as the original study. All hyperparameters were left at default to prevent any optimization based on the results of our previous study prediction GMA scores. As before, and the classifier was restricted to an ensemble size of one to minimize overfitting. This step ensured that model performance was consistent with previously published results and validated the robustness of the automated feature extraction and learning pipeline.

To extend this validation, we assessed whether the same model, previously trained to predict GMA scores, could generalize to CP outcomes without retraining. The classifier was originally trained to predict the “absent fidgety” movement type, a well-established early indicator of CP risk [13, 14, 31]. The model was pre-registered on May 22, 2024, before testing on the held-out lock-box dataset. It was downloaded on January 12, 2025, for evaluation on CP outcome data. The same 38-feature movement vector was re-computed to verify reproducibility, and the dataset was split into Train/Test/Holdout sets using pre-registered IDs. Positive class probabilities were predicted for the holdout set, and an ROC curve was constructed using CP outcomes as ground truth instead of GMA scores.

### 2.4 Cerebral Palsy Diagnosis and Outcome Prediction

Final diagnosis of cerebral palsy was determined through clinical assessment by pediatric physicians (pediatricians, physiatrists, neurologists) at CHOP following standardized diagnostic criteria [32]. These included persistent, nonprogressive motor impairments of neurological origin. Clinical evaluation of motor impairment was often supplemented by birth history and where available, neuroimaging findings (MRI) of brain lesion associated with CP. To ensure that the dataset used for outcome prediction was well-defined, only confirmed CP cases were included in the analysis. Among the 1,060 infants assessed using the GMA, 143 were later diagnosed with CP, with severity levels ranging from I (mild) to V (severe). All diagnosed CP cases were included in the dataset, regardless of severity or subtype.

Our goal was to determine whether the auto-sklearn model, trained exclusively on GMA scores, could successfully predict CP outcomes. By replacing GMA labels with confirmed CP diagnoses, we assessed the generalizability of early movement features in identifying eventual CP diagnosis. This provides an indication of whether movement-based biomarkers, initially optimized for GMA classification, also hold predictive value for long-term motor impairments.

## 3 Results

### 3.1 Replication of GMA Score Prediction from a Publicly Available Dataset

To confirm the reproducibility of our previous findings, we first validated that a model trained on movement-derived features using automated machine learning, without researcher-driven feature selection or hyperparameter tuning, could predict GMA scores with high accuracy. The original model achieved an AUC-ROC of 0.79 on a held-out test set [43]. To further ensure reproducibility, we trained a new model using the publicly released movement feature dataset [40] while incorporating updated versions of key Python libraries. The performance of this retrained model remained consistent with previous results, yielding an AUC-ROC of 0.78, indicating that the methodology is robust to minor variations in implementation.

### 3.2 Generalization of Model Predictions to CP Outcomes

We next assessed whether the same model trained to predict GMA scores could generalize to predicting CP outcomes without retraining. The classifier demonstrated minimal loss in performance, achieving an AUC-ROC of 0.74 when applied to CP diagnoses at long-term follow-ups. This result indicates that movement features originally used for GMA classification also capture clinically relevant signals that persist over time, supporting their use for direct neurodevelopmental outcome prediction.

To further evaluate the utility of this approach, we analyzed the model’s precision-recall trade-offs across different classification thresholds. When optimizing for a specificity-matched threshold (0.95) to align with GMA classification, the model exhibited lower sensitivity than GMA assessments (16% vs. 60%), indicating that a strict decision threshold may underdetect some high-risk cases. Conversely, when a threshold was chosen to maximize true positives while minimizing false negatives, the model’s true positive and false negative rates closely matched those observed in GMA classification. However, this threshold also resulted in a higher false positive rate, highlighting the trade-offs in sensitivity and specificity when using an automated model as a potential pre-screening tool.

We used a pre-registered lock-box test set for all final model evaluation, ensuring that CP outcome classification was performed on previously unseen data. This strict separation of training and testing data minimizes overfitting concerns and strengthens confidence in the model’s ability to generalize. Notably, the model more frequently misclassified less severe cases of CP as negative, while all severe CP cases were correctly classified as positive. This suggests that pronounced motor deficits are more easily detected by movement-based predictive models, whereas milder cases may require additional clinical context for accurate classification.

**Figure 2:**
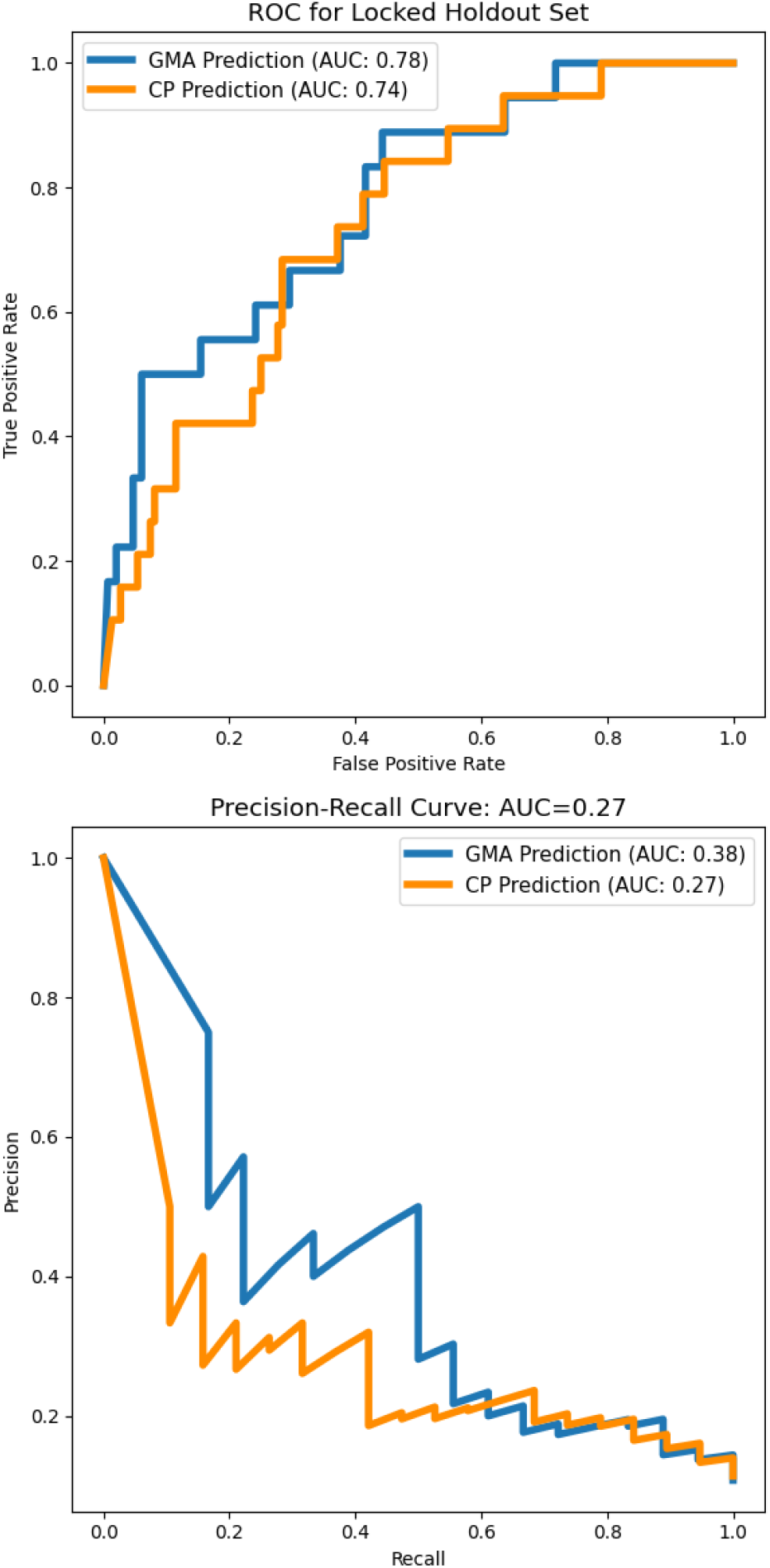
Model performance for GMA score and CP outcome prediction. (Top) ROC curves for the model trained to predict General Movement Assessment (GMA) scores, and evaluated on GMA score prediction (blue) and CP prediction (orange). The ROC curve for CP prediction shows an AUC of 0.74 indicating minimal loss in performance. (Bottom) PR curves for the same model, evaluated on GMA score prediction and CP outcome prediction without retraining. AUC for CP prediction (0.27) is lower than for GMA prediction (0.38) showing that some specificity is lost when generalizing to outcome prediction.

### 3.3 Comparison with Clinician Assessment

Both the model and clinicians successfully identified many infants who were later diagnosed with CP. However, there were some cases where both the model and gma score did not predict infants who were later diagnosed with CP (false negatives). Notably, both the model and clinicians did not correctly predict CP outcomes for a subset of 5 of the same infants, particularly those later diagnosed with mild CP (< III), and only one with a high severity (V), suggesting that the movement feature vector—designed to align with features clinicians use in GMA assessments—may not fully capture certain movement abnormalities. Additionally, some infants may only develop detectable deficits later in the neurodevelopmental trajectory, highlighting the need for longitudinal monitoring rather than a single timepoint-based assessment.

While AUC-ROC provides a global measure of classification performance, sensitivity and specificity are the clinically relevant metrics used in diagnostic decision-making. Sensitivity (*true positive rate*) quantifies the model’s ability to correctly identify infants who later develop CP, whereas specificity (*true negative rate*) measures its ability to correctly classify typically developing infants. These are defined as:

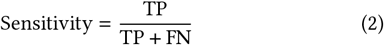

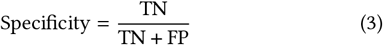

where TP (true positives) represents correctly identified CP cases, FN (false negatives) represents missed CP cases, TN (true negatives) represents correctly classified typically developing infants, and FP (false positives) represents misclassified healthy infants. These metrics are critical for determining the clinical utility of a screening tool, as high sensitivity minimizes missed diagnoses, while high specificity reduces unnecessary follow-up evaluations (see Table 1).

**Table 1:** Comparison of Model and Clinician Performance Metrics.

| Metric | Model (balanced) | Model (high specificity) | Clinician Prediction |
| --- | --- | --- | --- |
| Sensitivity | 0.68 | 0.16 | 0.58 |
| Specificity | 0.72 | 0.95 | 0.95 |

## 4 Discussion

### 4.1 Generalization of GMA-Based Predictions to CP Outcomes

This study evaluates whether a machine learning model trained to predict GMA scores can generalize to predict CP risk. Although the model was originally trained to predict GMA scores rather than CP outcomes directly, it still demonstrated strong predictive performance of risk.

Retraining the model specifically for the diagnosis of CP may yield even better results. However, the video dataset collected for this study was collected specifically to allow clinicians to evaluate the GMA, meaning that the infants were at rest and making only spontaneous movements. Therefore, a model trained directly on CP outcomes might struggle without incorporating additional clinical context beyond movement data alone.

Generalization without retraining is particularly significant because many machine learning models degrade in performance when applied to tasks they were not explicitly optimized for [12, 53**?**] and may be a sign of robustness. Unlike models that require fine-tuning on CP-labeled datasets, this approach shows promise in identifying long-term neurodevelopmental risk using features originally extracted for GMA classification. However, validation on external datasets, especially those collected prospectively as opposed to retrospectively, will be crucial to assess whether this generalization holds across different clinical settings and populations.

**Figure 3:**
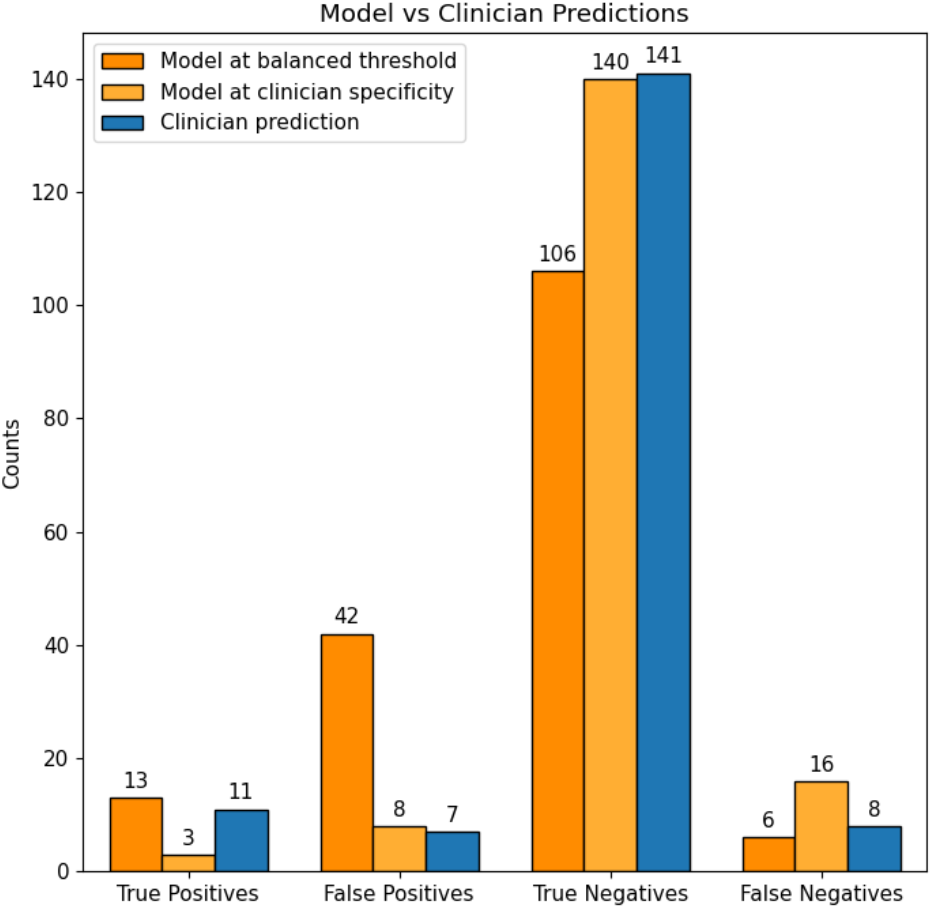
Sensitivity/Specificy tradeoff at different ROC thresholds. When selecting a threshold that balances True Positives and False Positives (dark orange), the model shows moderate sensitivity/specificity (0.68, 0.72), but a high proportion of False Positives relative to the clinician’s predictions indicated by GMA Score (blue). When selecting a threshold that matches clinician specificity of 95%, model has a much lower sensitivity of 0.16 (light orange)

### 4.2 Improving feature representations via self-supervised pre-training

The current approach relies on clinician-selected movement features, which are relatively coarse representations of infant motion. This was necessary to maintain consistency with prior work and assess the generalizability of the pre-registered model. However, hand-engineered features likely do not capture the full complexity of motor patterns, especially when averaged across all frames as was done in the present work. Future work will focus on learning movement features directly through masked pre-training approaches, which have been shown to improve feature extraction and model robustness in other domains [3, 28, 36, 37], and in movement analysis [4].

Deep learning-based models, particularly those using self-supervised learning techniques, have demonstrated remarkable success in capturing complex kinematic patterns across various domains [4, 9, 15, 23, 27]. Models pre-trained on large-scale adult datasets, including ViTPose-H [11], have already shown exceptional generalization capabilities in pose estimation and movement analysis [5]. Applying similar pretraining strategies to diverse infant movement datasets could unlock novel insights into early motor development, enhancing the ability to analyze small datasets that are prevalent in this field. By learning many of the features that describe human movement from large diverse datasets, deep learning approaches may overcome current limitations in dataset size for small, sepcialized populations like infants while preserving clinical interpretability. Future research should explore whether pretraining on large, heterogeneous datasets enables better feature extraction for infant movement analysis and improves predictive performance beyond manually selected kinematic descriptors.

### 4.3 Clinical Context and Potential for Integration

In reality, CP diagnosis is based on multiple factors beyond movement observation [32]. Clinicians integrate birth history, growth metrics, and standardized developmental assessments such as the Alberta Infant Motor Scale (AIMS) [25], Bayley Scales of Infant and Toddler Development [2], and Hammersmith Infant Neurological Examination [39]. They also rely on multiple observation sessions over time to track developmental progress. While this study, and many others demonstrates that movement-based models can contribute to early risk screening, future research should explore integrating broader clinical data to increase the sensitivity of the model while maintaining a high level of specificity.

Combining video-derived movement analytics with additional clinical variables could create a more comprehensive decision-support system. Multi-modal learning approaches that incorporate MRI findings, genetic predisposition, and caregiver-reported developmental concerns may enhance the robustness of automated screening tools as has been done in other fields of medicine [24]. Additionally, understanding how such models fit into existing clinical workflows will be critical for adoption, requiring discussions with healthcare providers to determine usability and transparency requirements.

### 4.4 Significance of Direct CP Outcome Prediction

The results show that a model trained to predict GMA scores (AUC-ROC 0.78) can also predict CP outcomes with minimal performance loss (AUC-ROC 0.74) without any retraining. This suggests that the movement features captured for GMA are also relevant for longterm neurodevelopmental outcomes. Unlike prior studies that stop at intermediate clinical scores, this evaluation directly tests CP outcomes. This distinction is critical because, although GMA is highly correlated with CP risk, it is not a perfect predictor. Some infants classified as high-risk by GMA develop typically, while others who were not flagged at an early stage later receive a CP diagnosis. A model explicitly trained to predict CP could potentially improve clinical utility by reducing false positives and false negatives.

This finding underscores the importance of moving beyond surrogate clinical scores when developing machine learning models for early diagnosis. By predicting clinical outcomes directly, models can provide more actionable insights for early intervention planning. However, future studies should consider the inherent uncertainty in long-term CP diagnoses and explore approaches for calibrating model confidence levels to account for developmental variability.

### 4.5 Reproducibility and Open Science in Clinical Machine Learning

The ability of this model to generalize without retraining highlights the importance of rigorous methodology in machine learning for clinical applications. The model was not optimized on the CP outcome labels, which were not available until 2+ years later. This underscores the importance of avoiding excessive feature engineering and tuning on a single dataset, which can lead to overfitting. Additionally, all code, data, and models from this study are preregistered and publicly available, allowing independent validation at other clinical sites. Open science improves reproducibility, and even if this model has limitations, making the findings transparent enables the research community to build better models in the future.

Reproducibility remains a challenge in medical AI research due to differences in hardware, dataset characteristics, and preprocessing pipelines across institutions [47]. Moreover, privacy constraints make it difficult share data across sites. By making models publicly available and open-sourcing all code and de-identified features, researchers can facilitate independent validation, identify potential biases, and ensure that models generalize beyond the datasets on which they were initially trained. Collaborative efforts involving federated learning and privacy-preserving techniques could further enable multi-site validation while respecting data privacy constraints, ultimately improving the robustness and clinical utility of AI-driven diagnostic tools.

### 4.6 Future Directions

Overall, these findings suggest that models trained to predict clinically relevant scores are likely learning features that capture subtle movement differences indicative of long-term outcomes. Future research will focus on improving feature extraction, refining model performance, and evaluating feasibility as both a global prescreening tool and a clinical decision-support system.

Next steps include increasing dataset diversity to evaluate generalizability across different infant populations, refining movement feature extraction methods using self-supervised learning, and integrating additional clinical variables to enhance predictive power.

## Data Availability

Data are available online at https://osf.io/bwgek/

https://zenodo.org/records/14674148

## Acknowledgments

This work was funded by an NIH-NICHD grant (Project#: 1R01HD097686, PIs: Johnson, Michelle J. and Kording, Konrad P.) and the clinical Early Detection Trial data collection was supported in part by the Cerebral Palsy Foundation.

